# Inhaled LTI-03 for Idiopathic Pulmonary Fibrosis: A Randomized Dose Escalation Study

**DOI:** 10.1101/2025.10.28.25338981

**Authors:** Philip L. Molyneaux, Nikhil A. Hirani, Collin C.K. Chia, Tejaswini Kulkarni, Tanzira Zaman, Robert J. Kaner, Ana Lucia Coelho, Yago Amigo Pinho Jannini-Sa, Brian Windsor, Sydney Kruger, Dale J. Christensen, Steven A. Shoemaker, Cory M. Hogaboam, BreAnne MacKenzie, Andreas Günther

**Affiliations:** Royal Brompton and Harefield Hospitals, Guy’s and St Thomas’ NHS Foundation Trust, London, United Kingdom; National Heart and Lung Institute, Imperial College London, London, United Kingdom; Edinburgh Lung Fibrosis Clinic, Royal Infirmary of Edinburgh, Edinburgh, United Kingdom; Institute for Regeneration and Repair, University of Edinburgh, Edinburgh, United Kingdom; Launceston Respiratory and Sleep Center, Launceston General Hospital, Launceston, TAS, Australia; Pulmonary, Allergy & Critical Care Medicine, University of Alabama Birmingham, Alabama, United States; Pulmonary & Critical Care Medicine, Cedars Sinai Medical Center, Los Angeles, California, United States; Weill Cornell Medicine, New York, New York, United States; Women’s Guild Lung Institute, Cedars Sinai Medical Center, Los Angeles, California, United States; Rein Therapeutics, Inc., Austin, Texas, United States; Department of Medicine, Duke University Medical School, Durham, North Carolina, United States; Center for Interstitial and Rare Lung Diseases, Justus-Liebig-University, University of Giessen and Marburg Lung Center, Member of the German Center for Lung Research, Giessen, Germany; Lung Clinic, Evangelisches Krankenhaus Mittelhessen, Giessen, Germany; Institute for Lung Health (ILH), Giessen, Germany; Excellence Cluster Cardiopulmonary Institute (CPI), Giessen, Germany

**Author notes:** **Corresponding Author**: Professor Andreas Günther, M.D. Center for Interstitial and Rare Lung Diseases, Justus-Liebig-University, Giessen University Hospital Giessen/Marburg, Klinikstrasse 36, 35392 Giessen, Germany.

## Abstract

Idiopathic pulmonary fibrosis (IPF) is a fatal interstitial lung disease with limited treatment options. LTI-03 promotes alveolar epithelial cell survival and reduces profibrotic protein expression in experimental models of IPF. In this Phase 1b, placebo-controlled, dose-escalation study, 24 participants with IPF were randomized 3:1 into 2 sequential dose cohorts to LTI-03 (5 or 10 mg/day) or placebo for 14 days (ClinicalTrials.gov: NCT05954988). The primary endpoint was the incidence of treatment-emergent adverse events (TEAEs). Exploratory analyses included pharmacokinetics and changes from baseline in expression of biomarkers related to fibrotic processes and epithelial integrity. Inhaled LTI-03 was well-tolerated, with no treatment-related discontinuations and only mild or moderate TEAEs; cough was the most common treatment-related TEAE. There was no evidence of airway obstruction by symptoms or spirometry. In bronchoscopy-derived deep bronchial brushing samples, both doses of LTI-03 significantly reduced interleukin-11 and thymic stromal lymphopoietin compared to placebo. Additionally, LTI-03 10 mg/day significantly reduced the expression of collagen type 1 alpha chain 1, CXC chemokine ligand 7 and galectin-7 compared to placebo. A trend in the reduction of plasma surfactant protein D was also observed in the LTI-03 10 mg/day group compared to placebo. The favorable safety and tolerability profile in addition to a reduction of disease-related biomarkers supports further evaluation of inhaled LTI-03 for IPF in a Phase 2 study (RENEW; ClinicalTrials.gov: NCT06968845).

## INTRODUCTION

Idiopathic pulmonary fibrosis (IPF) is a progressive, fatal, age-associated lung disease with poor survival^1^. The pathogenesis of IPF is characterized by apoptosis and senescence of alveolar epithelial type 2 cells, proliferation and accumulation of activated myofibroblasts, deposition of extracellular matrix (ECM), and fibrosis, resulting in progressive dyspnea and loss of lung function, especially forced vital capacity (FVC)^2,3^. The approved anti-fibrotic drugs nintedanib (Ofev^®^), pirfenidone (Esbriet^®^), and nerandomilast (Jascayd^®^) slow the rate of FVC decline^2,4^, but do not halt disease progression or restore lung function. Additionally, nintedanib and pirfenidone may be poorly tolerated due to gastrointestinal or cutaneous side effects, leading to frequent discontinuation^5,6^. Hence, there is an urgent need for therapies better targeting the underlying causes of IPF and other interstitial lung diseases; in particular, alveolar protective treatments that may act to restore healthy lung function with a more favorable tolerability profile. Caveolin-1 protein (Cav-1) is a structural protein that plays a critical role in regulating lung repair and cellular movement by restoring pathways that both promote epithelial health and halt fibrosis in the lung^7–9^. Cav-1 expression in pulmonary fibroblasts and alveolar epithelial cells is decreased in transgenic and rodent bleomycin models of lung injury and in the lungs of patients with IPF^10–12^. Augmentation of endogenous Cav-1 levels and supplementation of exogenous caveolin scaffolding domain (CSD) peptide have been shown to suppress lung fibrosis^7,10,11,13,14^. LTI-03 is a synthetic oligopeptide consisting of seven natural L-amino acids of the CSD. LTI-03 has been shown to exert strong antifibrotic and epithelial supportive effects in experimental animal models of lung fibrosis^15–17^, in human IPF precision cut lung slices^18^, and in human IPF alveolar organoids^19^.

While LTI-03 first demonstrated efficacy upon intraperitoneal administration in mouse models of IPF^20^, its limited aqueous solubility necessitated an alternate formulation and delivery strategy. Micronization of LTI-03 without the use of excipients demonstrated excellent stability, and importantly, the ability to achieve deep lung deposition^21^. LTI-03 (2.5 mg/capsule) is self-administered by study participants using a commercially available dry-powder inhaler. This study reports the safety and tolerability of LTI-03 following twice daily (BID) inhaled administration of LTI-03 (total doses of 5 mg/day or 10 mg/day) to participants with IPF for 14 days. Finally, LTI-03 treatment-related, impacts on exploratory biomarkers, which were selected based on nonclinical, translational experimental results, are presented and discussed.

## Results

### Participant Disposition and Characteristics

Twenty-four participants with IPF met eligibility criteria and were enrolled between June 2023 and October 2024 into 2 sequential dose cohorts: Cohort 1 (5 mg/day LTI-03 versus placebo), followed by Cohort 2 (LTI-03 10 mg/day versus placebo) (Figure 1). Within each cohort, participants were randomized 3:1 to study drug (LTI-03:placebo), which was self-administered BID for 14 days. No participant discontinued study treatment due to a treatment-emergent adverse event (TEAE), and all completed the study. LTI-03 dosing was incorrectly stopped by the Investigator on Day 14 for 1 participant in the LTI-03 5 mg/day group due to “met stopping criteria with decrease in forced expiratory volume in 1 second (FEV1)”; however, the protocol stopping criteria also required an associated decrease from baseline in the FEV1/FVC ratio, which was not met and would have supported continued dose administration. At baseline, demographics and disease characteristics were generally well balanced across the treatment groups (Table 1). No prior use of any approved or investigational antifibrotic drug was reported for any participant.

**Figure 1:**
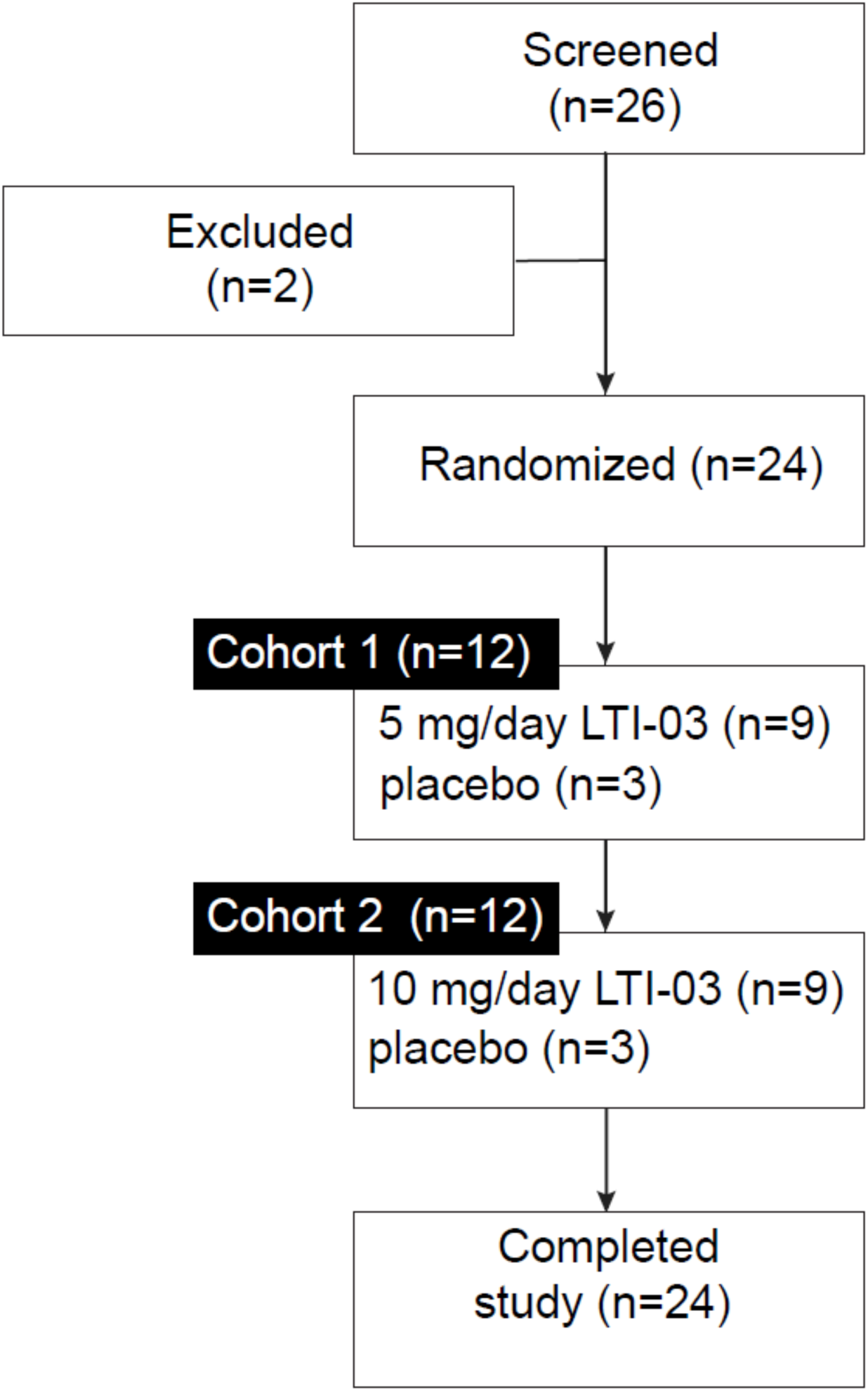
Phase 1b clinical study participant disposition

**Table 1.** Baseline Demographics and Disease Characteristics.

| Parameter | LTI-03 |  |  | Placebo |
| --- | --- | --- | --- | --- |
|  | 5 mg/day | 10 mg/day | Pooled | Pooled |
|  | (N = 9) | (N = 9) | (N = 18) | (N = 6) |
| Age, yr, mean (SD) | 67.8 (9.76) | 68.4 (10.79) | 68.1 (9.99) | 71.7 (6.15) |
| Male, *n* (%) | 7 (77.8) | 7 (77.8) | 14 (77.8) | 6 (100) |
| Female, *n* (%) | 2 (22.2) | 2 (22.2) | 4 (22.2) | 0 |
| Race and Ethnicity, *n* (%) |  |  |  |  |
| White, Not Hispanic or Latino | 9 (100) | 9 (100) | 18 (100) | 6 (100) |
| BMI (kg/m<sup>2</sup>), mean (SD) | 28.9 (2.73) | 29.4 (6.63) | 29.1 (4.93) | 29.3 (4.99) |
| Past/current cigarette use, *n* (%) | 8 (88.9) | 7 (77.8) | 15 (83.3) | 4 (66.7) |
| FVC percent predicted, mean (SD) | 84.8 (10.22) | 92.1 (18.66) | 88.5 (15.08) | 75.6 (8.94) |
| DLCO percent predicted, mean (SD) | 59.0 (7.66)* | 60.9 (11.70) | 60.0 (9.75)<sup>†</sup> | 61.4 (10.11) |
| LCQ total score, mean (SD) | 18.2 (3.26) | 16.6 (3.24) | 17.4 (3.26) | 16.1 (3.37) |
| Cough VAS score, mean (SD) | 29.6 (30.65) | 38.1 (21.51) | 33.8 (26.06) | 35.3 (27.20) |

*SD* standard deviation, *BMI* body mass index, *DLCO* diffusion capacity of the lungs for carbon monoxide, *FVC* forced vital capacity, *LCQ* Leicester Cough Questionnaire, *VAS* Visual Analog Scale.
\*Data available for n=8
<sup>†</sup>Data available for n=17

### Safety and Tolerability

LTI-03 was well tolerated over 14 days of BID dosing at total doses of 5 mg/day and 10 mg/day. The incidence of TEAEs was 66.7% (LTI-03 5 mg/day), 77.8% (LTI-03 10 mg/day), and 50% (placebo), respectively (Table 2). There were no fatalities or TEAEs leading to treatment discontinuation. One (4.2%) participant in the LTI-03 10 mg/day group experienced an unrelated serious TEAE of Grade 2 post-bronchoscopy fever requiring overnight inpatient observation. Most TEAEs in all dose groups were Grade 1 in severity. The total number of participants reporting Grade 2 TEAEs was low (n=3; 16.7%), and there were no TEAEs in any group that were Grade 3 or higher. Cough was the most common TEAE, reported by 3 (33.3%; LTI-03 5 mg/day), 5 (55.6%;

**Table 2.**
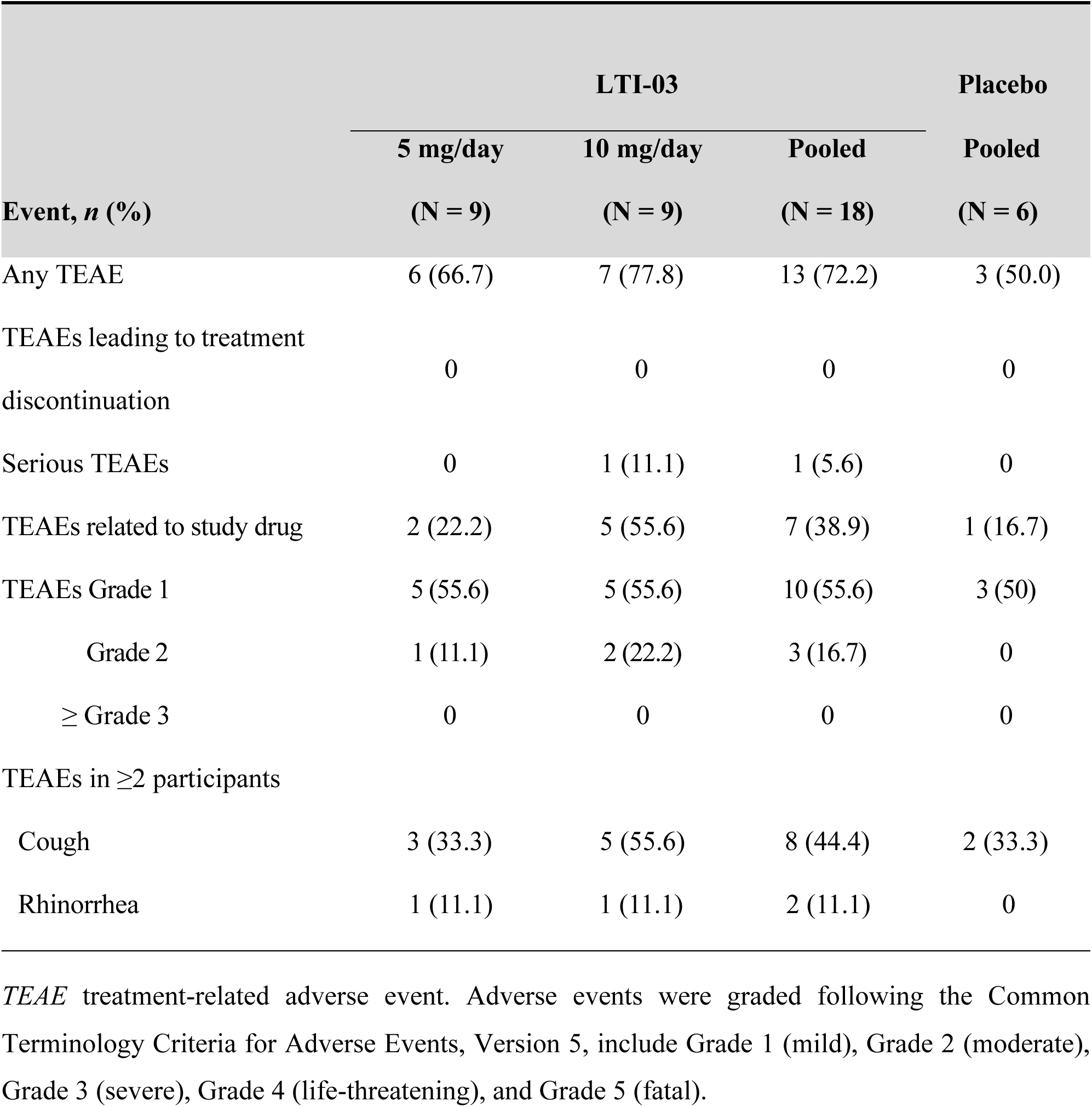
Overview of Treatment-Emergent Adverse Events.

|  | LTI-03 |  |  | Placebo |
| --- | --- | --- | --- | --- |
|  | 5 mg/day | 10 mg/day | Pooled | Pooled |
| Event, *n* (%) | (N = 9) | (N = 9) | (N = 18) | (N = 6) |
| Any TEAE | 6 (66.7) | 7 (77.8) | 13 (72.2) | 3 (50.0) |
| TEAEs leading to treatment discontinuation | 0 | 0 | 0 | 0 |
| Serious TEAEs | 0 | 1 (11.1) | 1 (5.6) | 0 |
| TEAEs related to study drug | 2 (22.2) | 5 (55.6) | 7 (38.9) | 1 (16.7) |
| TEAEs Grade 1 | 5 (55.6) | 5 (55.6) | 10 (55.6) | 3 (50) |
| Grade 2 | 1 (11.1) | 2 (22.2) | 3 (16.7) | 0 |
| ≥ Grade 3 | 0 | 0 | 0 | 0 |
| TEAEs in ≥2 participants |  |  |  |  |
| Cough | 3 (33.3) | 5 (55.6) | 8 (44.4) | 2 (33.3) |
| Rhinorrhea | 1 (11.1) | 1 (11.1) | 2 (11.1) | 0 |

*TEAE* treatment-related adverse event. Adverse events were graded following the Common Terminology Criteria for Adverse Events, Version 5, include Grade 1 (mild), Grade 2 (moderate), Grade 3 (severe), Grade 4 (life-threatening), and Grade 5 (fatal).

LTI-03 10 mg/day), and 2 (33.3%; placebo) participants, respectively. Rhinorrhea, reported by 1 participant in each of the LTI-03 treatment groups, was the only other TEAE reported by more than 1 participant in any group. A Grade 2 cough related to study drug that resolved on the same day was reported in 1 (11.1%) participant each in the LTI-03 5 mg/day and LTI-03 10 mg/day groups, respectively. Cough was the only study drug-related TEAE experienced by more than 1 participant. None of the study drug-related TEAEs was a serious TEAE or led to treatment discontinuation.

No apparent treatment- or dose-related trends in clinical laboratory parameters, vital sign measurements, 12-lead electrocardiogram (ECG) results, or physical examination findings were observed during the study. Mean changes from baseline in spirometry performed prior to and 30 and 60 minutes after in-clinic study drug administration were not clinically significant, indicating no acute effects of study drug inhalation on airways.

Participants self-reported cough symptoms using the Leicester Cough Questionnaire (LCQ)^22^ and a cough severity visual analogue scale (VAS). Baseline results are summarized in Table 1. No clinically significant changes in the LCQ total score or the cough VAS were observed during the treatment period for any treatment group, indicating that inhalation of study drug for 14 days did not appear to negatively affect participants’ quality of life related to cough or worsen pre-existing chronic cough symptoms.

### Pharmacokinetics

All post-dose plasma concentrations following LTI-03 inhalation were below the limit of quantification (BLQ) on Days 1, 7, and 14. LTI-03 concentrations in bronchoalveolar lavage fluid (BALF) at Day 14 were quantifiable in 3 participants in the LTI-03 5 mg/day group (1.55, 6.60, and 1.37 ng/mL) and 2 participants in the LTI-03 10 mg/day group (2.57 and 14.7 ng/mL); all other BALF sample concentrations were BLQ.

### Exploratory Biomarkers

Our exploratory biomarker strategy was based on the pathobiology of Cav-1 in the context of fibrosis^7,23–26^; IPF pathogenesis^27,28^, and importantly, pharmacodynamic data for LTI-03 from a variety of translational models^17–19^. Based on these data, it was hypothesized that LTI-03 treatment would cause a reduction of these biomarkers in a clinical setting. Plasma levels of surfactant protein D (SP-D), an indicator of epithelial cell stress, trended downwards and was decreased by 5% in the LTI-03 10 mg/day group compared to placebo. LTI-03 treatment did not change phosphorylated (p) AKT over total AKT levels in peripheral blood mononuclear cells (PBMCs). In deep bronchial brushing (DBB) samples, LTI-03 treatment significantly reduced expression of multiple profibrotic proteins compared to placebo, including collagen type 1 alpha chain 1 (COL1A1), chemokine (C-X-C motif) ligand 7 (CXCL7), galectin-7 (GAL-7), interleukin 11 (IL-11), and thymic stromal lymphopoietin (TSLP) (Table 3; Supplemental Figure 1).

**Table 3.**
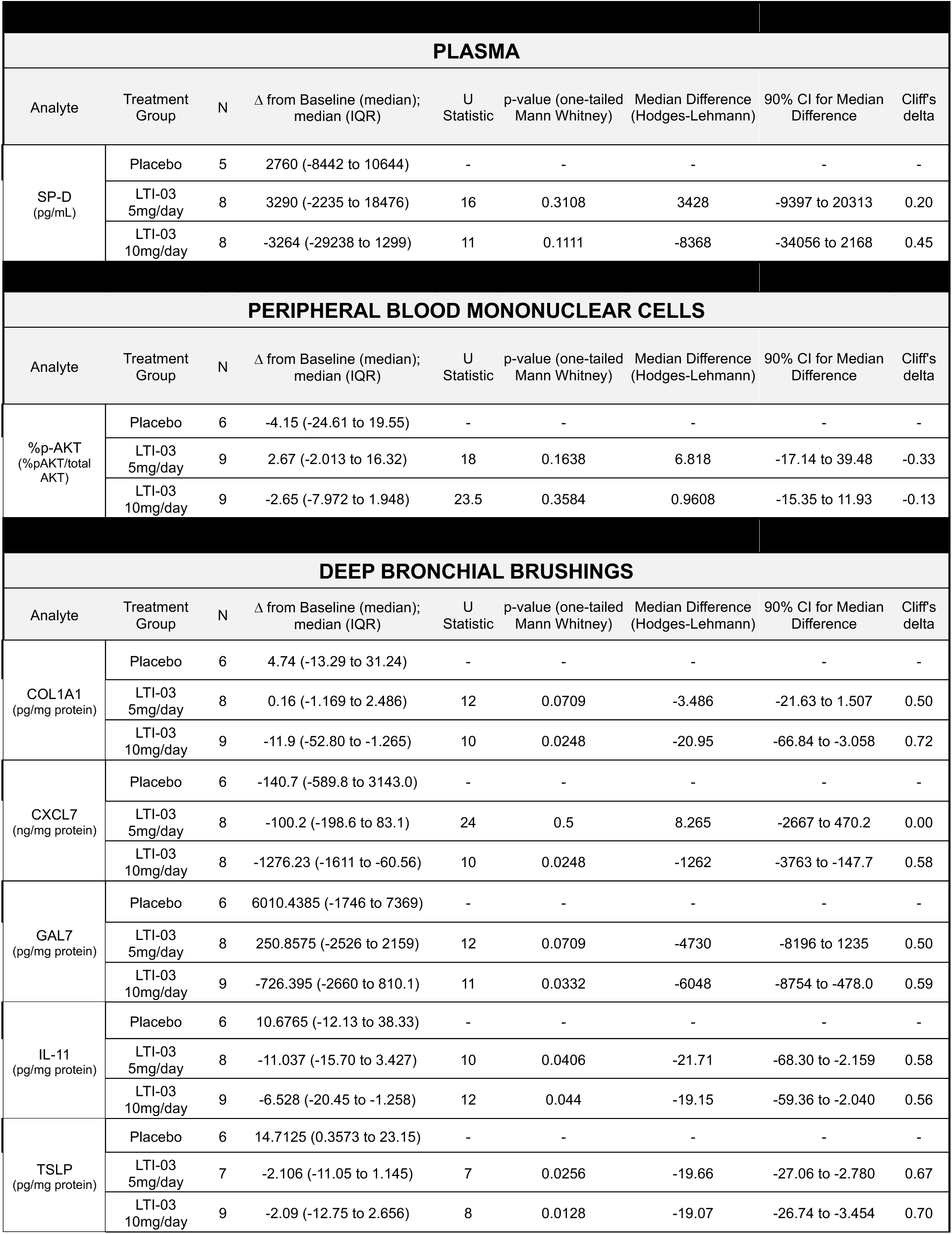
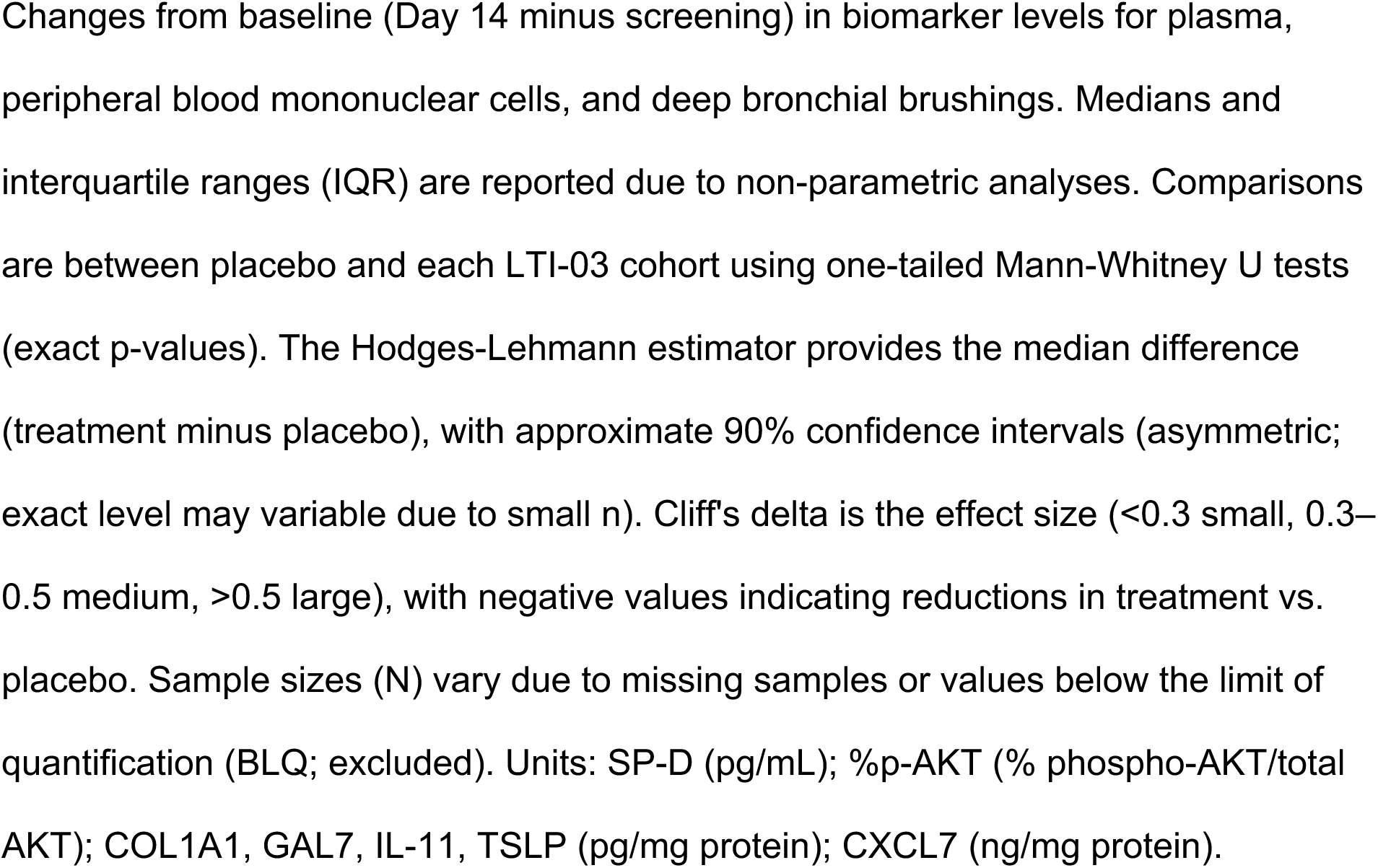
Exploratory Biomarkers Change from Baseline.

## Discussion

LTI-03 is a CSD peptide formulated as an excipient-free dry powder for inhalation. This first-in-patient study evaluated the safety and tolerability of LTI-03 compared to placebo when administered BID for 14 days in 24 participants with IPF. Participants were diagnosed with IPF within 3 years of screening per guidelines; none reported receiving prior treatment with nintedanib, pirfenidone, or nerandomilast. Inhaled LTI-03 at doses of 5 mg/day (2.5 mg BID) and 10 mg/day (5 mg BID) was well tolerated and all 24 participants completed the study. There were no TEAEs that led to treatment discontinuation. The majority of TEAEs were mild (Grade 1) in severity; only 3 (16.7%) participants reported Grade 2 AEs, and none (0%) reported TEAEs Grade 3 or higher. One participant in the LTI-03 5 mg/day group experienced a serious TEAE of Grade 2 fever following bronchoscopy that warranted overnight admission for observation; they had no clinical signs or symptoms of infection, no antibiotic treatment was required during or after hospital discharge, and the event was considered unrelated to study drug. Post-procedural fever is a known complication following bronchoscopy^29^.

Coughing is a hallmark of IPF and represents one of the key symptoms of IPF patients in larger cohorts^30^. In line with previous reports on inhaled drugs in IPF^31^, cough was also the most frequently reported TEAE for 44% of participants receiving LTI-03, as compared to the 33% receiving placebo, and the only study drug-related TEAE experienced by more than 1 participant. The majority of cough TEAEs were mild in severity except for 2 study drug-related TEAEs of Grade 2 severity, each resolving on the same day: one occurred as a single episode and the other occurred intermittently. All other study drug-related TEAEs of cough were intermittent in frequency and Grade 1 in severity, occurring intermittently after study drug administration. Other than rhinorrhea, which was reported for 1 participant each in the LTI-03 5 mg/day and 10 mg/day groups, no other TEAEs were reported by more than 1 participant.

Participants self-reported the effect of LTI-03 on their chronic cough symptoms and daily life using the LCQ^22^, and the severity of their cough symptoms using a visual analogue scale that rated cough severity (0 = no cough to 100 = worst cough ever). Self-reported LCQ and VAS results indicated that inhalation of the study drug did not appear to negatively affect participants’ quality of life or worsen pre-existing chronic cough symptoms.

Spirometry was performed pre- and post-dose (30 and 60 minutes) as a safety assessment to quantify any acute effects of study drug inhalation on airways. There were no clinically significant differences between the LTI-03 and placebo groups in mean changes in FEV_1_ and FVC immediately following study drug dosing and over the course of the treatment period. Furthermore, there was no evidence by either spirometry or participant-reported symptoms of airway obstruction following study drug inhalation.

Systemic levels of LTI-03 measured immediately after LTI-03 inhalation were undetectable in all participants regardless of LTI-03 dose, similar to healthy volunteers at the same doses (unpublished data). Canine toxicology studies have shown that this is likely due to absorption of LTI-03, a hydrophobic peptide, by lung tissue (unpublished data). Conversely, LTI-03 was detectable in BALF samples collected after inhalation for 5 participants (3 in the 5 mg/day; 2 in the 10 mg/day group), confirming distribution of LTI-03 into the lung. In BALF, the measurement of LTI-03 was limited by the low solubility of hydrophobic LTI-03 in aqueous solutions as well as differences in time between drug inhalation and sample collection via bronchoscopy and variability in the volume of BALF sample collected.

It is hypothesized that IPF pathophysiology is driven by the persistent microinjury of alveolar epithelial cells leading to apoptosis and necrosis, downstream aberrant fibroblast activation, and abnormal accumulation of extracellular matrix^3^. LTI-03 has been repeatedly shown to significantly attenuate profibrotic factors and pathways and also to support the maintenance of alveolar epithelial cell homeostasis in multiple animal models of fibrosis^17,20^, in precision cut lung slices (PCLS)^18^ that contain all diseased lung cell types, as well as in both lung epithelial cell organoid cultures^19^ and fibroblasts sourced from end-stage IPF patients^20^.

Exploratory biomarkers for this study were selected based on previous translational study outcomes which demonstrated robust, consistent reduction of these markers following LTI-03 treatment either *in vitro* or in *in vivo* animal models of IPF. Changes in biomarker levels from baseline to day 14 were assessed in plasma, PBMCs, and bronchoscopy-derived DBB samples. A one-tailed test was selected to test the directional hypotheses of reduced biomarker expression following LTI-03 treatment.

In plasma samples, levels of SP-D, an indicator of epithelial cell health, were decreased by 5% (p=0.1111) for the LTI-03 10 mg/day group compared to placebo. While not statistically significant, these reductions may be clinically relevant, as increased circulating SP-D levels are significantly linked to decline in lung function and are responsive to standard of care^32,33^.

Moreover, the 5% drop in SP-D is comparable to decreases described in standard of care IPF drug trials after longer treatment durations, notably the CAPACITY study of pirfenidone (5% at 12 weeks)^34^ and the INMARK study of nintedanib (4% at 12 weeks)^35,36^ which may indicate the potential for LTI-03 to slow the rate of lung function decline. In addition, SP-D is a key analyte in a platform currently in development by the PROLIFIC consortium, which is on track to gain regulatory approval for the creation of a diagnostic, prognostic, and theragnostic assay for IPF patients^37^.

While significant LTI-03 mediated decreases of p-AKT in IPF fibroblast monocultures have been observed (unpublished data), in this study LTI-03 compared to placebo did not modulate the percent phosphorylated AKT over total AKT in PBMCs from patients, suggesting that inhaled administration does not lead to changes in circulating immune cells.

In DBB samples, analyses revealed consistent reductions in key profibrotic and inflammatory markers with LTI-03 compared to placebo, particularly at the 10 mg/day dose level. LTI-03 consistently reduced collagen expression and deposition in animal models and in vitro^17,20^. Levels of COL1A1, a marker of extracellular matrix deposition, decreased significantly with LTI-03 10 mg/day compared to placebo.

Similarly, CXCL7, a chemokine involved in inflammatory processes in the lung, was significantly attenuated in DBB samples from participants who received LTI-03 10 mg/day. These results are consistent with prior data demonstrating decreased CXCL7 levels with LTI-03 treatment in IPF PCLS supernatants^18^. CXCL7 levels have been reported to be elevated in bronchoalveolar lavage fluid (BALF) from patients with IPF compared with healthy controls^38^. While chemokine signaling is increasingly recognized as contributing to IPF pathobiology, the specific role of CXCL7 in disease initiation or progression has not yet been defined^39^.

Galectin-7, a protein associated with epithelial repair and fibrosis^17^ was reduced for both the LTI-03 5 mg/day and 10 mg/day groups compared to placebo. Immunohistochemical analyses of IPF lung tissue have shown intense GAL-7 expression localized to the alveolar and bronchiolar epithelium and alveolar septum^12^. The LTI-03-mediated reductions in GAL-7 in DBB samples suggest that inhaled LTI-03 had a targeted effect in the deep lung.

Interleukin-11 levels in DBB samples were significantly reduced in both LTI-03 treatment groups compared to placebo. This is consistent with results from aged male mice challenged with bleomycin, in which BALF levels of IL-11 were reduced with LTI-03 treatment^17^. IL-11 is a key driver of myofibroblast activation and differentiation in IPF^40^, and is currently the focus of an ongoing clinical study in IPF with an investigational monoclonal antibody (ClinicalTrials.gov: NCT07036523).

Robust reductions of TSLP were also observed in both the LTI-03 5 mg/day and 10 mg/day treatment groups compared to placebo. TSLP is an alarmin cytokine that is released following epithelium damage and promotes type 2 inflammation and fibrosis, driving pathological processes such as mucus hypersecretion and airway remodeling^41^. Levels of TSLP are significantly elevated in BALF^42^ and lung tissue from patients with IPF^43^ and immunohistochemical studies have localized elevated TSLP expression to alveolar epithelial cells and fibrotic foci^43^. In IPF, TSLP may promote fibrogenesis by activating fibroblasts and recruiting immune cells to sites of epithelial injury^43^. Statistically significant reductions in TSLP following LTI-03 treatment support the hypothesis that LTI-03 may have positive effects on lung epithelial cell health.

The effects of LTI-03 treatment on DBB biomarkers suggest attenuation of fibrotic pathways and support of epithelial integrity in the lung microenvironment, aligning with LTI-03’s mechanism of action targeting caveolin-1-mediated signaling. Despite limitations of small sample sizes, these data support disease-modifying potential of LTI-03, warranting further clinical evaluation with functional endpoints to confirm disease modification in IPF.

There remains an unmet medical need for safe and effective treatments that target epithelial cell injury in IPF. Inhaled therapies can facilitate increased drug delivery to the site of disease while minimizing systemic off-target effects. The overall low incidence of serious, treatment-related, or severe TEAEs, absence of treatment discontinuations due to a TEAE, and no detectable systemic absorption suggest that inhaled LTI-03 has the potential for a more favorable safety, tolerability, and adherence profile compared to the currently approved, systemically administered treatments. Preclinical data in experimental models and evidence of active LTI-03 pharmacodynamics in this study suggest that LTI-03 may regulate processes driving both lung repair and fibrosis.

Limitations in the interpretation of this study include the small sample size (n=24) and a short treatment duration of 14 days. A sample size of 24 participants was planned to ensure that safety and tolerability of LTI-03 were adequately assessed while minimizing unnecessary participant exposure. No conclusion of LTI-03 effects on disease progression can be derived from the data collected. Larger studies of longer treatment durations in patients with IPF will be required to confirm and expand upon the findings of this first-in-patient study.

This study showed that the administration of inhaled LTI-03 to participants with IPF was safe and well-tolerated at doses of 5 and 10 mg/day. Additionally, an exploratory analysis of pro-fibrotic, epithelial health, and inflammatory biomarkers suggests a potential pharmacodynamic benefit of LTI-03 administration without effects on circulating immune cells. These encouraging data support further evaluation of inhaled LTI-03 in a Phase 2 study of longer duration in patients with IPF (RENEW; ClinicalTrials.gov: NCT06968845).

## METHODS

### Study Design

This was a Phase 1b, randomized, double-blind, placebo-controlled, dose escalation study conducted at 6 sites in the United States, United Kingdom, Germany, and Australia (ClinicalTrials.gov: NCT05954988). The primary objective was to assess the safety and tolerability of inhaled LTI-03 compared to placebo in patients with IPF. Exploratory objectives were to assess LTI-03 pharmacokinetics (PK) and pharmacodynamics.

The study had 3 periods: screening (up to 21 days), treatment (14 days) and follow-up (7 days), with study visits on Days 1, 7, 14, and 21 (Figure 2). The protocol is provided as a supplement.

**Figure 2:**
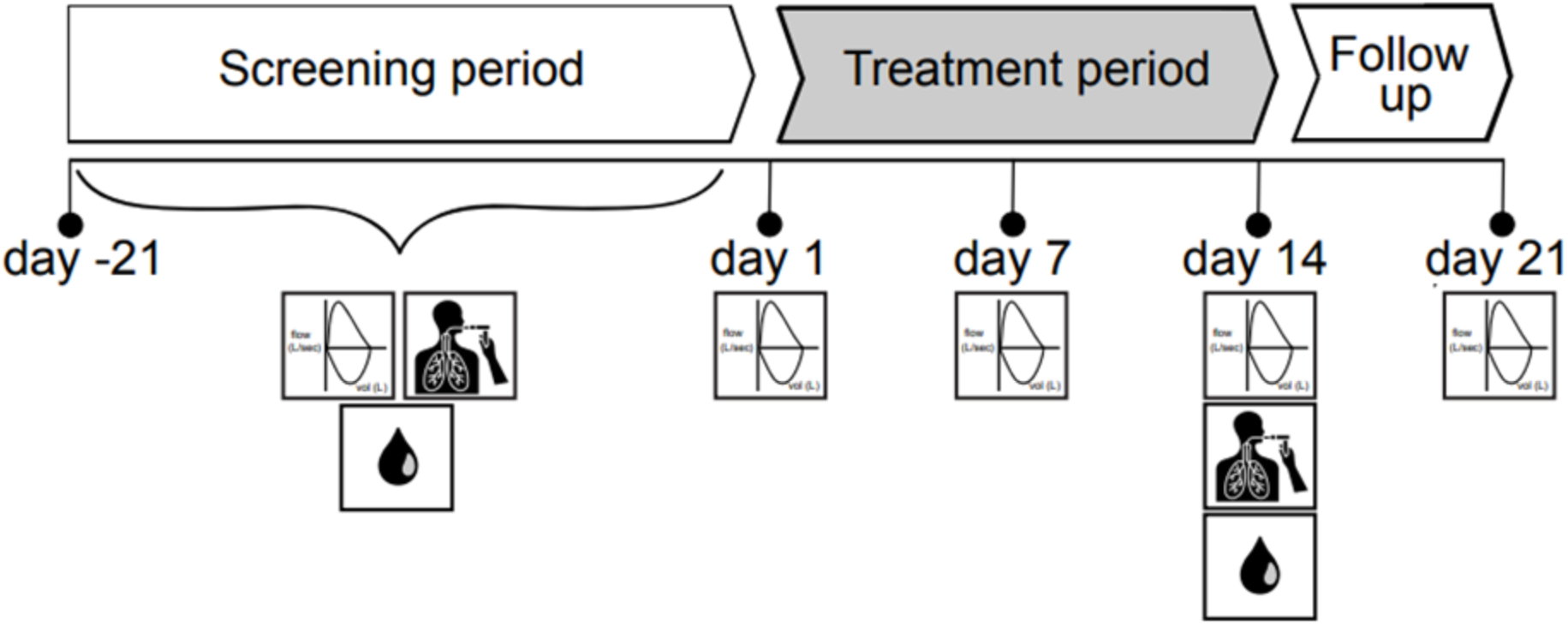
Phase 1b clinical study protocol design. Images reflect spirometry, bronchoscopy, and blood biomarker collection timepoints. Blood for biomarker analyses were collected during screening prior to the bronchoscopy.

### Study Population

Participants were recruited between 22 June 2023 and 01 October 2024. Eligible participants were ≥ 40 years of age with a diagnosis of IPF per ATS/ERS/JRS/ALAT guidelines^44^, made within 3 years of enrollment, with FVC percent predicted ≥ 40%, DLCO percent predicted between 30% and 80%, and FEV₁/FVC ≥ 0.7. Participants with interstitial lung disease other than IPF, significant obstructive lung disease, current diagnosis of asthma, or who experienced a pulmonary exacerbation within 6 months prior to screening were excluded, as were those with significant renal or hepatic impairment. In addition, participant could not have received treatment with an approved or investigational anti-fibrotic therapy for IPF within 2 months of the baseline bronchoscopy.

### Randomization Treatment and Blinding

Eligible participants were randomized 3:1 to LTI-03 or placebo into 2 sequential cohorts (n=12 per cohort). The 5 mg/day (low-dose) cohort received 2.5 mg LTI-03 or placebo BID and the 10 mg/day (high dose) cohort received 5 mg BID LTI-03 or placebo. Randomization was performed by interactive response system. Study staff were blinded to treatment assignment throughout the study.

The study complied with Good Clinical Practice and the Declaration of Helsinki. All participants provided written informed consent. The study protocol and informed consent were approved by each site’s Institutional Review Board or Independent Ethics Committee.

### Study Procedures

Safety and tolerability assessments were performed through treatment and follow-up and included evaluation of TEAEs, concomitant medications, physical examinations, vital signs, 12-lead ECGs, clinical laboratory assessments, spirometry, and cough symptoms using the validated LCQ^22^ and a cough VAS. Spirometry was performed according to American Thoracic Society (ATS) / European Respiratory Society (ERS) Task Force guidelines^45^ at all clinic visits, with assessments conducted both pre-and post-dose on study drug dosing days. Study staff monitored participants’ self-administration of study drug (the morning dose) during on-site visits and symptoms were monitored for at least 1 hour.

### Bronchoscopy Sample Collection

Samples collected by bronchoscopy were used to assess LTI-03 lung concentrations and to evaluate pharmacodynamic biomarkers of inhaled LTI-03. Bronchoscopy was performed to collect DBB samples, BALF, and/or bronchoabsorption (BA) samples at baseline and then on Day 14, approximately 2-3 hours after the participant self-administered their morning dose of study drug (LTI-03 or placebo) and after all other study assessments had been completed.

Deep bronchial brushings were required for all participants; the site had the option to collect either BAL or BA samples, or both, depending on local expertise, training, and procedural confidence. The sequence of procedures was to be BA, BAL, and then brushings to minimize the risk of blood contamination of samples. Sedation, local anesthesia, and post-procedure monitoring were performed according to the investigative site’s standard of care. Either oral or nasal bronchoscopy routes were acceptable.

Given significant variations in volume of BAL fluid retrieved and that only two sites successfully collected bronchoabsorption samples, biomarker data presented describe results from DBB samples only.

### Deep Bronchial Brushings

Bronchoscopists were instructed to obtain a total of 6 brushings from 3 of the basilar bronchial segments of the right lower lobe: the anterior basilar segment, the lateral basilar segment, and the posterior basilar segment. With the bronchoscope proximal to or within the orifice of the appropriate basilar segment, the brush sheath was to be passed out 5 cm beyond the subsegment orifice, and then the brush fully extended an additional 1 cm for a total of 6 cm beyond the orifice of the subsegmental bronchus. At the desired depth, the sheath was to be gently pulled back and forward for 15 strokes to brush the small airways. The brushing process was repeated using any combination of the 3 basilar segments described above, at the discretion of the bronchoscopist, for a total of 6 six separate brush samples.

### Bronchoalveolar lavage

Bronchoscopists were instructed to lavage a total of 150 mL of normal saline, in 30-50 mL aliquots, to either of the two bronchial segments of the right middle lobe: the lateral segment and the medial segment. If the airways were too small to access the medial or lateral segments, then the bronchoscope could be wedged in the right middle lobe bronchus. With the bronchoscope wedged in the appropriate right middle lobe segment (or right middle lobe bronchus if necessary), 30-50 mL of room temperature or warmed normal saline was injected using an appropriately sized syringe (e.g., 50-60 ml syringe) via tubing attached to the working channel of the bronchoscope lumen. Using the same syringe, the saline was aspirated out through the lumen. Wall suction into a sterile trap could be used as an alternative to syringe aspiration.

Samples for measurement of LTI-03 lung concentrations were collected from the first aspirate.

### Pharmacokinetic Analysis

Blood samples for the measurement of systemic LTI-03 concentrations were collected pre-dose (trough) and +5 minutes after dosing on study Days 1, 7, and 14.

### Biomarker Analysis

Blood samples for biomarker assessments in PBMCs and platelet rich plasma were collected prior to initial dose and on day 14 following last dose. Following optimization for detection of analytes in IPF matrices, meso-scale discovery electrochemiluminescence (MSD-ECL) assays were performed at BioAgylitix for SP-D (#K1519XR-2, MSD) in plasma and AKT/pAKT (#K15100D-1, MSD) in PBMCs according to manufacturer’s instructions.

Protein isolation and preparation of DBB samples for ELISAs were performed as previously described^46^ according to manufacturer’s instructions for COL1A1 (#ab210966, abcam), CXCL7 (#LS-F4967-1, LS Bio), GAL7 (#DY1339, R&D Systems), IL-11 (#DY218, R&D Systems), TSLP (#DY1398, R&D Systems).

### Statistical Analyses

All descriptive statistical analyses on measurements other than biomarkers were performed using SAS statistical software (Version 9.4). Missing data were not imputed unless otherwise stated. Continuous data were summarized using the mean, SD, median, minimum value, and maximum value. Categorical data were summarized using frequency counts and percentages.

The primary endpoint was the incidence of TEAEs reported from Day 1 through follow-up. TEAEs were coded using MedDRA (Version 26.1) and graded using the National Cancer Institute Common Terminology for Adverse Events (CTCAE) Version 5. Safety analyses included all participants who received study drug. A sample size of 18 LTI-03 participants provided a 60% chance of detecting an AE with a true incidence rate of 5% and an 85% chance of detecting a more common AE with a true incidence rate of 10%. Placebo participants in each cohort were pooled for safety and biomarker analyses. No secondary endpoints were specified.

Since exploratory biomarkers were selected based on data generated by prior translational experiments which demonstrated reduction following LTI-03 treatment in various models, one-tailed statistical analyses were performed. Sample results for placebo-treated participants from each cohort were pooled to increase power (from n=3 to n=6). Due to small sample sizes (n=5 to 9 per group) and non-normality confirmed by Shapiro-Wilk tests (p<0.05 for TSLP, IL-11, COL1A1, SP-D), a one-tailed Mann-Whitney U test (exact method) was used to compare changes from baseline to Day 14 between the pooled placebo group and each LTI-03 treatment group (5 and 10 mg/day).

Due to errors in either sample collection (leading to missing samples) or results below the lower limit of detection, there was occasional variability in total number of samples. For statistical analysis of plasma SP-D, blood samples were inadvertently not collected for the following participants: one in the placebo group (baseline and Day 14 missed), one in the LTI-03 5 mg/day (baseline missed), and a participant in LTI-03 10 mg/day (baseline and Day 14 missed), reducing the total sample size to N=5 for placebo and N=8 for each LTI-03 treatment group. Due to a COVID-19 infection, DBB samples were not collected for one participant in the LTI-03 5 mg/day treatment group on Day 14, reducing the total sample size to N=8 for analyses of all DBB biomarkers in this group. For TSLP, one result was BLQ from one participant in the LTI-03 5 mg/kg treatment group and the data point was excluded, reducing the sample size for the statistical analysis of TSLP to N=7.

Descriptive statistics are reported as medians with interquartile ranges (IQR). The Hodges-Lehmann estimator was used for the median difference between groups (treatment minus placebo), with approximate 90% confidence intervals (asymmetric, exact level varied slightly with small n). Cliff’s delta was calculated as effect size using the formula δ = (2U / (n1 × n2)) - 1, with thresholds: |δ| <0.3 small, 0.3–0.5 medium, >0.5 large. The sign was flipped to negative when treatment changes were lower than placebo (indicating reductions). Analyses were performed using GraphPad Prism version v10.0.

### Data sharing statement

Rein Therapeutics, Inc. (“Rein”) understands and acknowledges the need to share clinical study data with the research community in an open and transparent manner and has provided de-identified patient data in the manuscript. Source data are provided for biomarker analyses. Rein will not consider further requests pertaining to clinical data outside of what has been accepted and published by the journal. Any queries regarding clinical study data must be submitted in writing to.

## Data Availability

All data produced in the present work are contained in the manuscript.

## Acknowledgements

The authors would like to thank the participants, investigators, and their teams who took part in this study. The authors also acknowledge Walker Clinical Sciences, LLC for manuscript preparation assistance.

## Contributions

All authors contributed substantially to this study. Study Concept and design: P.L.M., N.A.H., C.K.C., T.K., T.Z., R.J.K., S.A.S., S.K., A.G. Acquisition, analysis, interpretation of the data: C.M.H., B.M., D.J.C., S.A.S., S.K. A.L.C., YA.P.J., B.W. Review and approval of the manuscript: P.L.M., N.A.H., C.K.C., T.K., T.Z., R.J.K., S.A.S., S.K., A.L.C., YA.P.J., C.M.H., B.M., D.J.C., B.W., A.G.

---

**Supplemental Figure 1.**
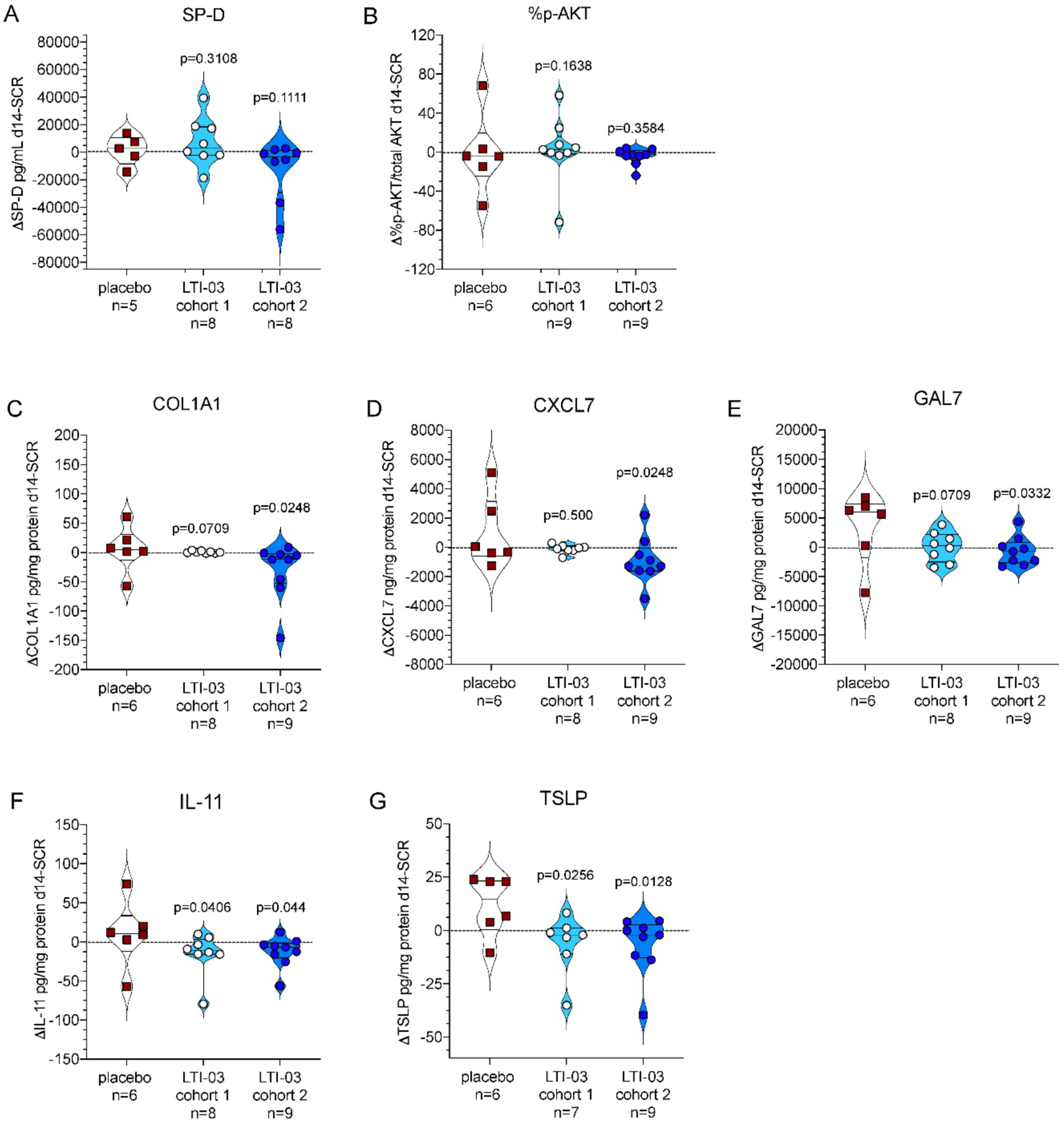
Exploratory biomarkers, change from baseline compared to placebo. Violin pots showing interquartile ranges with individual analyte change in baseline values overlaid are shown. One-tailed, unpaired, non-parametric Mann Whitney U tests were performed on analyte changes from baseline (pre-dose) to Day 14 for placebo (pooled) vs. LTI-03 5 mg/day or LTI-03 10 mg/day. A p-value of ≤ 0.05 was considered statistically significant. N: total number of samples. Data points were excluded either due to missed sample collection or an analyte measurement being below lower limit of quantitation (BLQ). Units: SP-D (pg/mL in plasma); %p-AKT (% phospho-AKT/total AKT in PBMCs); COL1A1, CXCL7, GAL7, IL-11, TSLP (pg/mg protein) or ng/mg protein for CXCL7. See Table 3 for full statistics, including effect sizes and confidence intervals. No multiple comparison correction applied (exploratory analysis). DBB = deep bronchial brushings; SCR = screening (baseline).

